# Machine learning for analysis of active stand responses in older adults with vasovagal syncope

**DOI:** 10.1101/2020.12.07.20241109

**Authors:** Michelle Kwok, Hugh Nolan, Chie Wei Fan, Clodagh O’Dwyer, Rose A Kenny, Ciarán Finucane

## Abstract

**Objectives:** To assess 1) differences in the hemodynamic response to the active stand test in older adults with a clinical diagnosis of vasovagal syncope compared to age-matched controls 2) if the active stand test combined with machine learning approaches can be used to identify the presence of vasovagal syncope in older adults.

**Approach:** Adults aged 50 and over (Vasovagal Syncope N=46 Age=66.9±10.3; Control N=86 Age=65.3±9.5) completed an active stand test. Multiple features were extracted to characterize the hemodynamic responses to the active stand test and were compared between groups. Classification was performed using machine learning algorithms including linear discriminant analysis, quadratic discriminant analysis, support vector machine and an ensemble majority vote classifier.

**Main Results:** Subjects with vasovagal syncope demonstrated a higher resting (supine) heart rate (69.8±13.1 bpm vs 63.3±12.1 bpm; P=0.007), a smaller initial systolic blood pressure drop (−20.2±20.1% vs −27.3±17.5%; P=0.005), larger drops in stroke volume (−14.7±24.0% vs −2.7±23.3%; P=0.010) and cardiac output (−6.4±18.5% vs 5.8±22.3%;P<0.001) and a larger increase in total peripheral resistance (8.1±30.4% vs −6.03±22.8%; P=0.002) compared to controls. A majority vote classifier identified the presence of vasovagal syncope with 82.6% sensitivity, 76.8% specificity, and average accuracy of 78.9%.

**Significance:** Older adults with vasovagal syncope display a unique hemodynamic and autonomic response to active standing characterized by relative autonomic hypersensitivity and larger drops in cardiac output compared to age-matched controls. With suitable machine learning algorithms, the active stand test holds the potential to be used to screen older adults for reflex syncopes and hypotensive susceptibility potentially reducing test time, cost, and patient discomfort. More broadly this paper presents a machine learning framework to support use of the active stand test for classification of clinical outcomes of interest.

## 1. Introduction

The head up tilt test (HUTT), (Kenny et al. in 1986), is used to confirm the diagnosis of reflex syncopes, such as vasovagal syncope (VVS). More recently it has been considered a test of hypotensive susceptibility in patients with a history of syncope (Moya et al. 2009). The test provokes a syncopal event in a controlled setting. However, it has moderate repeatability, is uncomfortable for patients, is difficult to perform in the very young or older frailer patients, is resource intensive, and is restricted to specialized clinics (Matsushima, Tanaka, and Tamai 2004; Benditt et al. 1996).

Previous research has explored methods to shorten the duration of the HUTT through early prediction algorithms or by replacing the test with an alternative. The best performing algorithms have been developed by Mereu et al. (2013), Stewart et al.(1996), Ebden (2006), and Virag et al. (2007; 2018), which combine parameters of heart rate (HR), blood pressure (BP), and heart rate variability (HRV) measured during the pre-syncopal stages of a HUTT for VVS prediction, achieving an accuracy of 86% to 94% (Virag et al. 2007; 2018; Mereu et al. 2013; Stewart, Erb, and Sorbera 1996; Ebden 2006). However, these algorithms still require significant time (up to 40 minutes) undergoing a HUTT and have not focussed on identifying patients with a clinical diagnosis of VVS.

Carmody et al. (2014) investigated the use of the active stand (AS) test for identification of those with a clinical diagnosis of VVS (as compared to HUTT outcomes) and achieved promising results in a young cohort with a multivariate classifier achieving a sensitivity of 84.3%, specificity of 72.9%, and accuracy of 80.2%. However, it was hypothesized that these results may not generalise well to all ages, since it is well known that the AS test and HUTT responses vary according to age. Older adults tend to exhibit blunted hemodynamic responses to orthostatic stress compared to age matched norms (Folino et al. 2010; Kurbaan et al. 2003; Laitinen et al. 2004; O’Dwyer 2011; Imholz et al. 1990; Dambrink and Wieling 1987; van Wijnen et al. 2017), which would be at odds with previous work suggesting a relative hypersensitivity in VVS in younger adults (Carmody et al. 2014). Most prediction algorithms, with the exception of Ebden (2006) and Virag et al. (2007; 2018), have not focussed on the older adult population, while only Carmody et al [8] has focussed on identifying a clinical diagnosis of VVS.

This retrospective cross-sectional study aims to address this gap by investigating the hypothesis that older adults with a clinical diagnosis of VVS exhibit multivariate hemodynamic differences during the AS test as compared to age-matched controls (CON) that can be used to predict a positive clinical diagnosis of VVS using machine-learning approaches.

## 2. Methods

### 2.1 Subjects

A retrospective analysis of VVS cases and controls (CON) aged 50 and over was collated from: 1) VVS patient data from the Falls and Syncope Unit (FASU) at St James’s Hospital Dublin (O’ Dwyer et al, 2011); and 2) CON data from the Irish Longitudinal Study on Ageing (TILDA); the contributions from each source are summarized in Figure 1. Approval for this study was obtained from the local research ethics committee.

**Figure 1.**
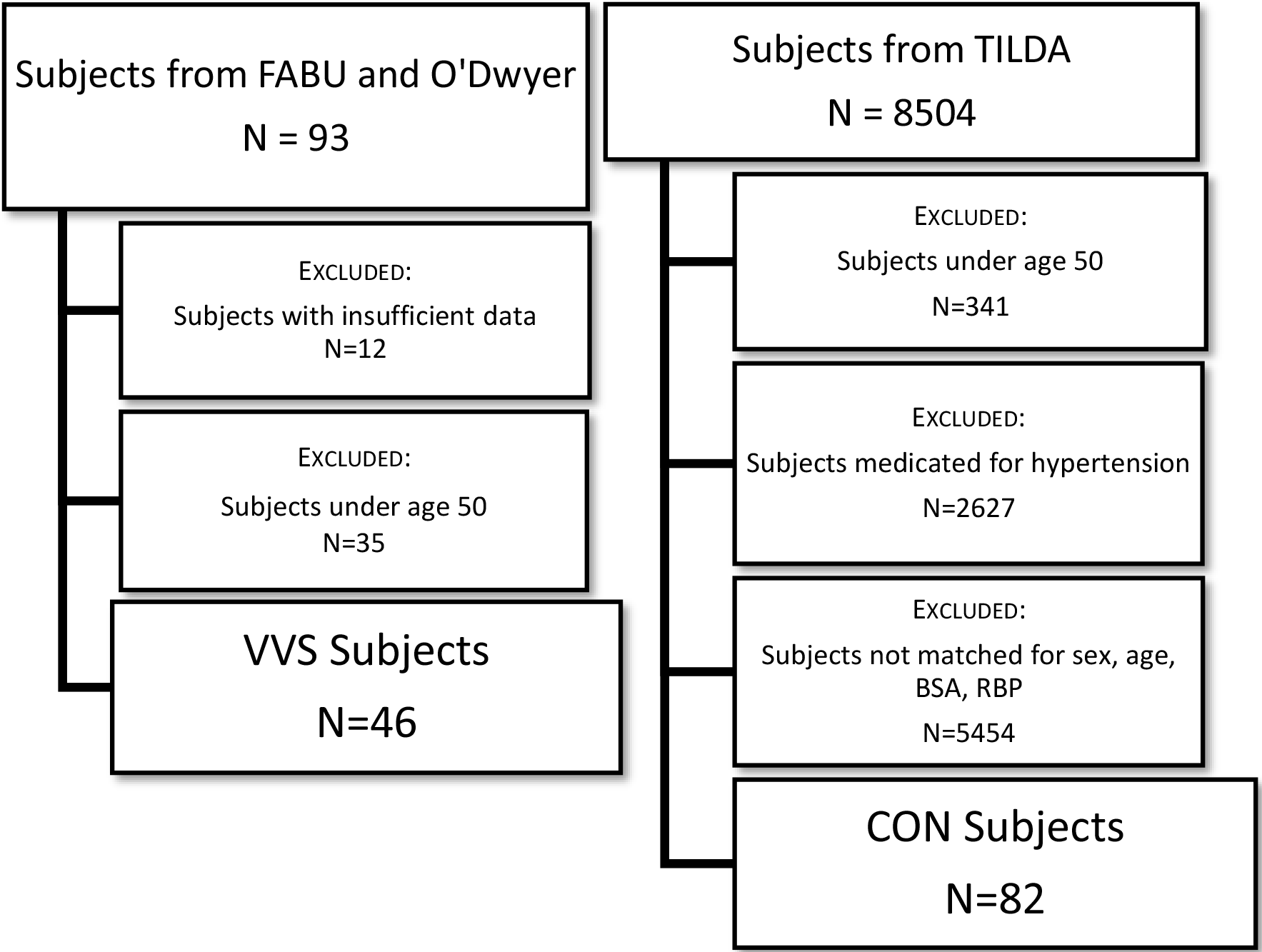
Summary of data utilized in study

VVS cases were recruited from the FASU between August 2008 and December 2014 (O’Dwyer 2011). This group included patients clinically evaluated (by HUTT), and reviewed by two independent, consultant geriatricians, who had a history consistent with VVS (denoted VVS+) (i.e. syncope precipitated by prolonged standing, fear, severe pain, emotional distress and associated with prodromal symptoms). Patients that were diagnosed with a form of syncope other than VVS were excluded. All patients in these groups had completed an AS test. The cause of syncope was based on history, and exclusion of other known causes of syncope in accordance with the European Society of Cardiology’s (ESC) guidelines.

A sample of CON subjects aged 50 and over were selected from TILDA who had completed an AS test as part of a health assessment and had no self-reported history of falls, unexplained falls or faints, and were not on medications for hypertension. These subjects did not however complete a HUTT or clinical assessment to confirm absence of diagnosis of syncope. CON subjects were matched to the VVS cases based on age, sex, body surface area (BSA), and resting BP. Matching between VVS subjects and CON subjects was confirmed using a two-tailed t-test on the matched parameters.

### 2.2 Experimental Protocol

Data collected for each VVS and CON subject included the responses of hemodynamic parameters generated during an AS test. The test involved 10 minutes of supine rest followed by 2 minutes of upright standing in a quiet, temperature controlled (21 to 23 °C), dimly lit room. This protocol is in line with the current protocol for the AS test where cardiovascular responses are seen within the first 2 to 3 minutes of standing (Finucane et al. 2019). Continuous measurements of systolic blood pressure (SBP), diastolic blood pressure (DBP), mean blood pressure (MBP), heart rate (HR), cardiac output (CO), stroke volume (SV), and total peripheral resistance (TPR) responses were collected using the Finometer® (200 Hz sampling frequency; 12 bit A/D). BP and HR measurement were obtained using the volume clamp method for measuring continuous finger arterial pressure and CO, SV, and TPR were estimated using the Modelflow method (Jellema 2005).

### 2.3 Analysis

The steps involved in this analysis are outlined in Figure 2. Data analysis was completed using MATLAB v17.0 by MathWorks (MathWorks, 3 Apple Hill Drive, Natick, MA 01760-2098, United States). Text files from the Finometer® were imported to MATLAB and interpolated from a beat-to-beat to a second-by-second scale to ensure comparability across individuals. The signal from the Finometer height correction unit data was used to identify the start-time of the stand.

**Figure 2.**
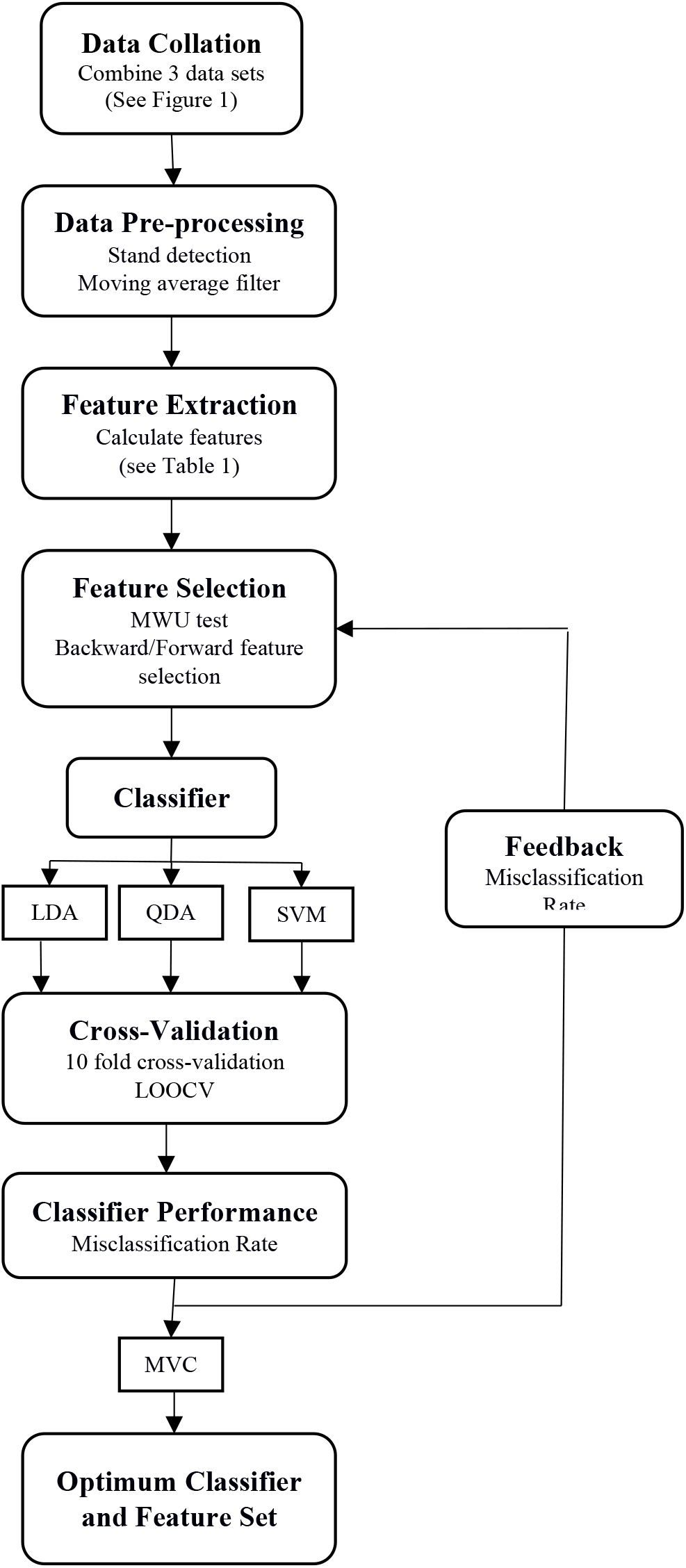
Summary of analysis. LDA = Linear discriminant analysis; QDA = Quadratic discriminant analysis; SVM = Support vector machine; MVC = Majority vote classifier; LOOCV = Leave one out cross-validation; MWU = Mann Whitney U Test

**Figure 3.**
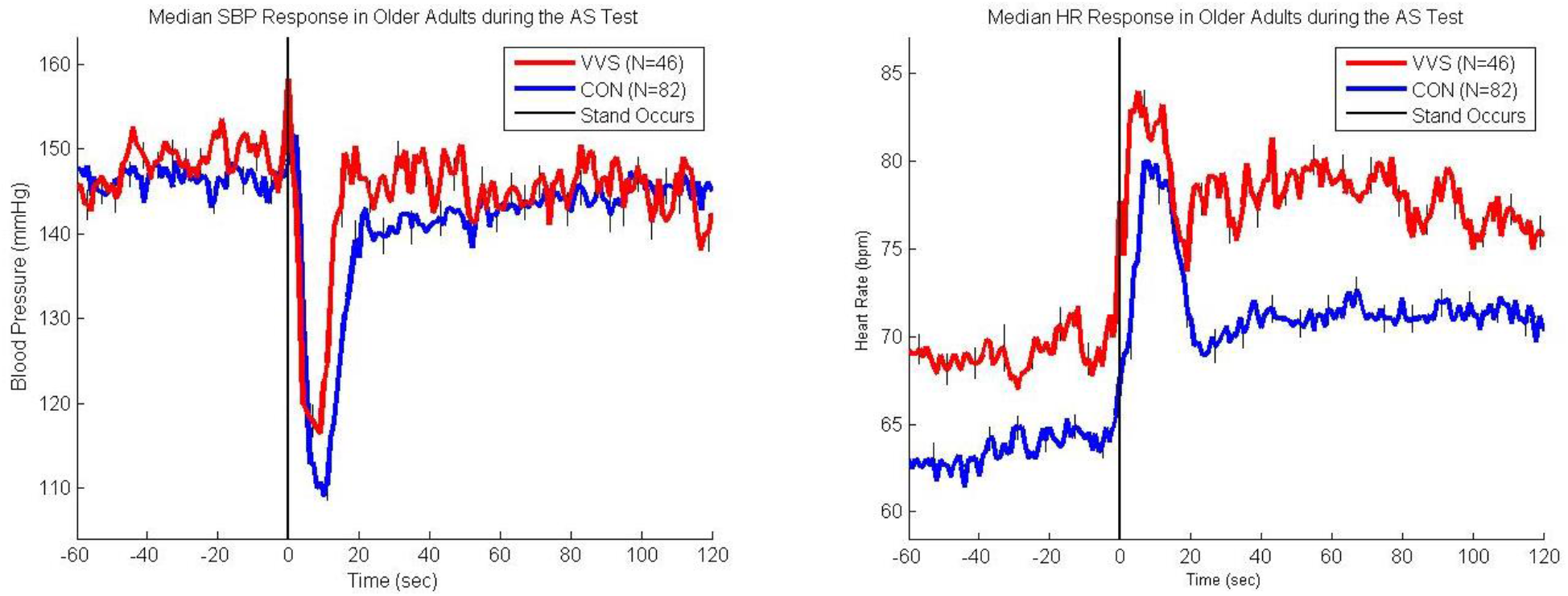
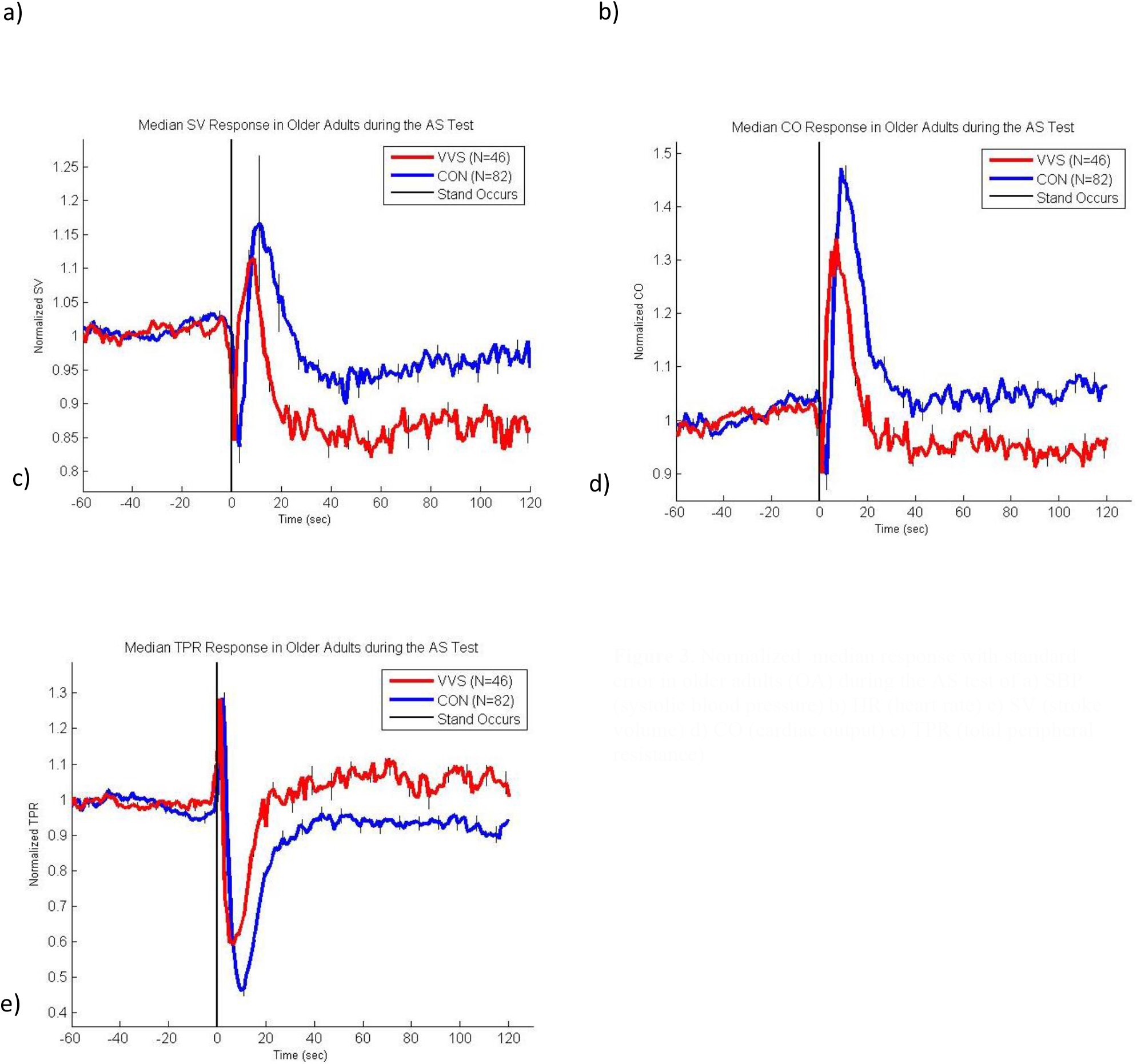
Normalized median response with standard error in older adults (OA) during the AS test of a) SBP (systolic blood pressure) b) HR (heart rate) c) SV (stroke volume) d) CO (cardiac output) e) TPR (total peripheral resistance)

### 2.4 Feature Extraction

The median response of the following signals were compared between older adult VVS and CON subjects: SBP, HR, rate-pressure product (RPP), SV (normalized by baseline mean), CO (normalized by baseline mean), TPR (normalized by baseline mean), heart rate variability (HRV) and blood pressure variability (BPV). Features of each signal were extracted based on trends previously identified in younger adults by Carmody et al. (Carmody et al. 2014) and newly identified trends to account for age-related differences in AS responses. Values of baseline and steady-state mean, trough and peak width, percentage increase and decrease from baseline, percentage change from baseline to steady-state, and the steady-state slope were calculated from each hemodynamic response signal resulting in a total of 72 candidate features. Baseline and steady-state values were defined as 60 to 30 seconds pre stand and 80 to 120 seconds post stand, respectively. These parameters and their method of computation are summarized in Table 1. For the HRV and BPV signals, total power in low (0.04 Hz to 0.15 Hz) and high frequency (0.15 Hz to 0.4 Hz) bands were computed at baseline and during standing (i.e. steady-state). The ratio of these values, along with the root mean square and standard deviation of successive differences were computed.

**Table 1.** Summary of parameters

| Parameter | Abbreviation | Units | Definition |
| --- | --- | --- | --- |
| Baseline mean | B | - | $B = \frac{1}{50} \sum_{i=-60}^{-10} x_i$ |
| Steady state mean | SS | - | $SS = \frac{1}{40} \sum_{i=80}^{120} x_i$ |
| Trough width | TW | S | $TW = i_{PK+} - i_{PK-}$<br>$i_{TR}$ is index of first trough > 0s<br>$i_{PK+}$ is index of first peak > $i_{TR}$<br>$i_{PK-}$ is index of last peak < $i_{TR}$ |
| Peak width | PW | S | $PW = i_{TR+} - i_{TR-}$<br>$i_{PK}$ is index of first trough > 0s<br>$i_{TR+}$ is index of first peak > $i_{PK}$<br>$i_{TR-}$ is index of last peak < $i_{PK}$ |
| Percentage increase from baseline | %I | % | $\%I = \left( \frac{Max(x_i) - Baseline}{Baseline} \right) \times 100$ |
| Percentage decrease from baseline | %D | % | $\%D = \left( \frac{Min(x_i) - Baseline}{Baseline} \right) \times 100$ |
| Percentage difference from baseline to steady state | B2SS | % | $B2SS = \left( \frac{SteadyState - Baseline}{Baseline} \right) \times 100$ |
| Steady state slope | SSS | - | $p(x) = p_1 x$ where $p_1$ is the SSS |
| Baseline low frequency | BLF | mV<sup>2</sup>/Hz | $BLF = \sum_{i=0.04}^{0.15} p_{xx}$ between -10s to -60s |
| Steady state low frequency | SLF | mV<sup>2</sup>/Hz | $SLF = \sum_{i=0.04}^{0.15} p_{xx}$ between 80s to 120s |
| Baseline high frequency | BHF | mV<sup>2</sup>/Hz | $BHF = \sum_{i=0.15}^{0.4} p_{xx}$ between -10s to -60s |
| Steady state high frequency | SHF | mV<sup>2</sup>/Hz | $SHF = \sum_{i=0.15}^{0.4} p_{xx}$ between 80s to 120s |
| Ratio of baseline low to high frequency | BLF/BHF | Ratio | $\frac{BLF}{BHF} = \frac{\sum_{i=0.04}^{0.15} p_{xx}}{\sum_{i=0.15}^{0.4} p_{xx}}$ between -10s to -60s |
| Ratio of steady state low to high frequency | SLF/SHF | Ratio | $\frac{SLF}{SHF} = \frac{\sum_{i=0.04}^{0.15} p_{xx}}{\sum_{i=0.15}^{0.4} p_{xx}}$ between 80s to 120s |
| Ratio of baseline to steady state low frequency | BLF/SLF | Ratio | $\frac{BLF}{SLF} = \frac{\sum_{i=0.04}^{0.15} p_{xx} \text{ between } -10s \text{ to } -60s}{\sum_{i=0.04}^{0.15} p_{xx} \text{ between } 80s \text{ to } 120s}$ |
| Ratio of baseline to steady state high frequency | BHF/SHF | Ratio | $\frac{BHF}{SHF} = \frac{\sum_{i=0.15}^{0.4} p_{xx} \text{ between } -10s \text{ to } -60s}{\sum_{i=0.15}^{0.4} p_{xx} \text{ between } 80s \text{ to } 120s}$ |
| Ratio of baseline low frequency to steady state high frequency | BLF/SHF | Ratio | $\frac{BLF}{SHF} = \frac{\sum_{i=0.04}^{0.15} p_{xx} \text{ between } -10s \text{ to } -60s}{\sum_{i=0.15}^{0.4} p_{xx} \text{ between } 80s \text{ to } 120s}$ |
| Ratio of steady state low frequency to baseline high frequency | SLF/BHF | Ratio | $\frac{SLF}{BHF} = \frac{\sum_{i=0.04}^{0.15} p_{xx} \text{ between } 80s \text{ to } 120s}{\sum_{i=0.15}^{0.4} p_{xx} \text{ between } -10s \text{ to } -60s}$ |
| Baseline root mean square | BRMS | S | $BRMS = \sqrt{\frac{1}{N-1} \sum_{i=1}^{N-1} ((R-R)_{i+1} - (R-R)_i)^2}$<br>Where N=number of pulse intervals (PI) |
| Steady state root mean square | SSRMS | S | $SSRMS = \sqrt{\frac{1}{N-1} \sum_{i=1}^{N-1} ((R-R)_{i+1} - (R-R)_i)^2}$<br>Where N=number of pulse intervals (PI) |
| Baseline standard deviation of successive differences | BSD | S | $BSD = \sqrt{\left( \frac{\sum_{i=1}^{N-1} (D_i - D_{mean})^2}{N-1} \right)}$ $D_1 = RR_1 - RR_2$ $D_{N-1} = RR_{N-1} - RR_N$ |
| Steady state standard deviation of successive differences | SSSD | S | $SSSD = \sqrt{\left( \frac{\sum_{i=1}^{N-1} (D_i - D_{mean})^2}{N-1} \right)}$ $D_1 = RR_1 - RR_2$ $D_{N-1} = RR_{N-1} - RR_N$ |

#### 2.4.1 Feature subset selection

A Mann-Whitney U (MWU) test was used to rank and identify important features that showed significant differences between VVS and CON subjects in order to reduce the number of features used. A p-value ≤0.05 was considered significant. Once significant candidate features had been identified, forward and backward sequential feature selection was applied to determine optimal feature sets based on minimization of the misclassification rate:

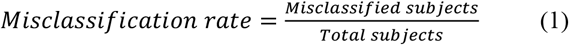

Forward and backward selection were each performed three times, once for each of three classification algorithms: linear and quadratic discriminant analysis (LDA and QDA) and a support vector machine (SVM).

### 2.5 Classifiers

The performance of three classification algorithms were compared. An LDA classifier was first tested following the success of Carmody et al. (2014) in classifying VVS in younger adults. This approach creates a discriminant function that generates a linear boundary between group distributions that minimises the misclassification rate. Future samples are classified based on the relative position of these features to this linear boundary.

In the LDA, the covariance (dispersion) matrix is assumed equal across groups. This assumption is most accurate when samples display approximate Gaussian distribution that can be easily separated by a linear boundary (Krzanowski 2000). Since it was observed that not all features could be separated by a linear boundary and were non-Gaussian, a QDA classification algorithm was also explored. In this classifier, the discriminant function takes on the form of a quadratic function (Krzanowski 2000).

A third classifier was tested using a SVM, which uses more complex methods to separate data of different classes. This algorithm performs classification based on the use of hyperplanes created in higher dimensional spaces and is often used to classify samples that are not linearly separable (Cristianini 2000).

Finally, to improve results further, an aggregating majority vote classifier was tested that combined the outputs of the three optimal classifiers by assigning the class ruled by the majority of the three classification algorithms.

#### 2.5.1 Classifier performance

Cross-validation was performed to evaluate classifier performance for the LDA, QDA, SVM, and majority vote classifier. An N-fold scheme was used that randomly segregates the data into N equally sized datasets which are subsequently used for training of the classifier (N-1 sets) and testing the classifier performance (1 set). This process is repeated based on the number of folds specified and results averaged across all trials. For this study, a 10-fold cross-validation scheme and leave-one-out cross-validation (LOOCV) scheme were used. The method uses the same process as the N-fold scheme but the number of folds is equivalent to the number of samples.

Classifier performance was evaluated using sensitivity, specificity, positive predictive value (PPV), negative predictive value (NPV), and accuracy. These measures were calculated, as follows:

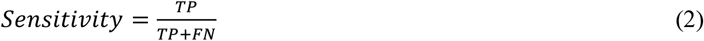

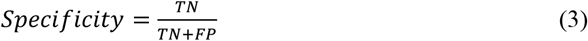

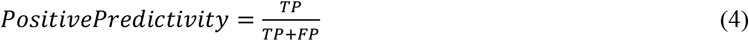

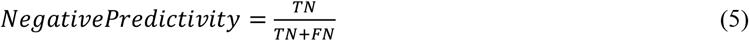

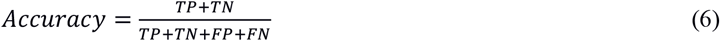

based on the number of true positives (TP), true negatives (TN), false positives (FP), and false negatives (FN).

## 3. Results

### 3.1 Subject Demographics

Subject characteristics used to match controls and subjects are detailed in Table 2.

**Table 2:** Summary of characteristics of matched older adult subjects and controls

|  | Units | VVS<br>N=46 | CON<br>N=82 | P-Value |
| --- | --- | --- | --- | --- |
| Age | Years (Mean ± SD) | 66.9 ± 10.3 | 65.3 ± 9.5 | 0.387 |
| Number of Males | N | 33 | 34 | 0.861 |
| Height | cm (mean ± SD) | 166.7 ± 10.6 | 164.3 ± 7.9 | 0.180 |
| Weight | Kg (mean ± SD) | 74.9 ± 15.4 | 74.9 ± 14.4 | 0.997 |
| Body surface area | m<sup>2</sup> (mean ± SD) | 1.9 ± 0.23 | 1.8 ± 0.20 | 0.744 |
| Resting BP | mmHg (mean ± SD) | 149.3 ± 20.1 | 145.7 ± 16.9 | 0.281 |

### 3.2 Differences in the Hemodynamic Response

#### 3.2.1 VVS vs. CON

Twenty-three parameters showed significant differences between VVS and CON. These results are summarized in Table 3. Subjects with VVS demonstrated a smaller drop in SBP (SBP-10s), a larger change in baseline to steady-state values in SV, CO and TPR (SV-B2SS, CO-B2SS, TPR-B2SS), and a smaller drop in TPR compared to CON subjects (TPR-%D). VVS subjects had a higher HR throughout the test (HR-B/HR-SS).

**Table 3.** Median and inter-quartile range for each derived parameter with MWU significance for VVS and CON subjects

|  |  | VVS | CON | VVS vs. CON |
| --- | --- | --- | --- | --- |
| Parameter | Units | Median (IQR) | Median (IQR) | P-Value |
| SBP-10s | % | -20.2 (20.1) | -27.3 (17.5) | 0.005 |
| HR-B | Bpm | 69.8 (13.1) | 63.3 (12.1) | 0.007 |
| HR-SS | Bpm | 76.5 (15.6) | 70.9 (15.7) | 0.026 |
| HR-%I | % | 31.7 (19.3) | 43.6 (38.9) | 0.012 |
| RPP-B | mmHg·bpm | 1.0e+04 (2.8e+03) | 9.5e+03 (2.0e+03) | 0.012 |
| SV-%I | % | 26.8 (36.9) | 36.0 (41.2) | 0.048 |
| SV-B2SS | % | -14.7 (24.0) | -2.7 (23.3) | 0.010 |
| SV-SSS | ml/sec | -0.031 (0.22) | 0.06 (0.14) | 0.022 |
| CO-%I | % | 54.2 (40.1) | 71.4 (48.5) | 0.018 |
| CO-B2SS | % | -6.4 (18.5) | 5.8 (22.3) | <0.001 |
| CO-SSS | l/min | -1.1e-3 (18e-3) | 1.6e-3 (9.6e-3) | 0.039 |
| TPR-%D | % | -46.8 (15.1) | -57.8 (17.7) | <0.001 |
| TPR-B2SS | % | 8.1 (30.4) | -6.03 (22.8) | 0.002 |
| HRV-BLF | mV<sup>2</sup>/Hz | 6.0e-3 (0.01) | 0.01 (0.03) | 0.007 |
| HRV-BLF/BHF | Ratio | 1.6 (2.9) | 3.0 (4.0) | 0.036 |
| HRV-SLF/SHF | Ratio | 3.4 (7.1) | 1.6 (2.6) | 0.016 |
| HRV-BLF/SLF | Ratio | 0.57 (2.5) | 2.6 (5.9) | <0.001 |
| HRV-SLF/BHF | Ratio | 2.8 (7.4) | 1.0 (2.2) | 0.010 |
| HRV-BRMS | S | 0.9 (0.2) | 0.9 (0.2) | 0.006 |
| HRV-SRMS | S | 0.8 (0.2) | 0.8 (0.2) | 0.024 |
| BPV-SLF | mV<sup>2</sup>/Hz | 509.3 (812.1) | 298.8 (531.0) | 0.021 |
| BPV-SLF/SHF | Ratio | 4.3 (10.0) | 2.9 (4.1) | 0.035 |
| BPV-BLF/SLF | Ratio | 0.5 (0.9) | 1.0 (2.2) | 0.009 |

VVS = Subjects with diagnosis of vasovagal syncope
CON = Control subjects
SBP = Systolic blood pressure
HR = Heart rate
RPP = Rate pressure product
SV = Stroke volume
CO = Cardiac output
TPR = Total peripheral resistance
HRV = Heart rate variability
BPV = Blood pressure variability
B = Baseline
SS = Steady state
10s% = 10 s percentage from baseline
%I = Percentage increase from baseline

The baseline low frequency (BLF) HRV values were also significantly greater in VVS. As a result, ratios to the baseline high frequency (HRV-BLF/BHF) and steady-state low and high frequency (HRV-BLF/SLF, HRV-BLF/SHF) also were significantly different. In the BPV signal, the median SLF was significantly higher in VVS subjects leading to significant differences in the BPV ratios BPV-SLF/SHF and BPV-SLF/BLF in VVS and CON older adults.

### 3.3 VVS Classification

#### 3.3.1 Feature selection

The feature sets identified using forward and backward sequential feature selection for the LDA, QDA, and SVM classifiers are as follows:

- LDA: RPP-B; CO-B2SS; HRV-BLF/SLF; HRV-SSRMS; BPV-SLF/SHF
- QDA: SBP-10s%; HR-SS; RPP-B; SV-B2SS; CO-B2SS; TPR-%D; TPR-B2SS; HRV-BLF/BHF; HRV-BLF/SLF; HRV-BRMS; HRV-SSRMS; BPV-SLF/SHF; BPV-BLF/SLF
- SVM: SBP-10s%; HR-SS; CO-B2SS; HRV-BLF/BHF; HRV-BLF/SLF; HRV-SLF/BHF; BPV-SLF/SHF

#### 3.3.2 Classification results

Results for cross-validation of the majority vote classifier and each of the constituent classifiers are summarized in Table 4. SVM (79.2%) and QDA (72.7%) classifiers outperform LDA classifiers (70%). The majority vote classifier successfully improved overall classifier performance by 0.8-3.1% depending on the cross-validation scheme utilised. A maximum accuracy of 78.9% was achieved with the majority vote classifier when LOOCV was performed.

**Table 4:** VVS vs. CON older adults multivariate classifier cross-validation (k-fold and LOOCV)

| 2-Fold Cross-Validation |  |  |  |  |  |
| --- | --- | --- | --- | --- | --- |
|  | Sensitivity | Specificity | PPV | NPV | Accuracy |
| **LDA** | 0.398 | 0.824 | 0.550 | 0.716 | 0.672 |
| **QDA** | 0.620 | 0.740 | 0.563 | 0.793 | 0.711 |
| **SVM** | 0.657 | 0.740 | 0.582 | 0.799 | 0.719 |
| **Majority Vote** | 0.690 | 0.761 | 0.667 | 0.782 | 0.734 |

| 5-Fold Cross-Validation |  |  |  |  |  |
| --- | --- | --- | --- | --- | --- |
|  | Sensitivity | Specificity | PPV | NPV | Accuracy |
| LDA | 0.598 | 0.861 | 0.755 | 0.779 | 0.760 |
| QDA | 0.768 | 0.784 | 0.677 | 0.852 | 0.776 |
| SVM | 0.871 | 0.630 | 0.587 | 0.893 | 0.720 |
| Majority Vote | 0.615 | 0.822 | 0.660 | 0.816 | 0.752 |
| 10-Fold Cross-Validation |  |  |  |  |  |
|  | Sensitivity | Specificity | PPV | NPV | Accuracy |
| LDA | 0.460 | 0.816 | 0.615 | 0.742 | 0.700 |
| QDA | 0.635 | 0.748 | 0.626 | 0.802 | 0.717 |
| SVM | 0.870 | 0.750 | 0.680 | 0.908 | 0.792 |
| Majority Vote | 0.763 | 0.853 | 0.745 | 0.852 | 0.800 |
| LOOCV |  |  |  |  |  |
|  | Sensitivity | Specificity | PPV | NPV | Accuracy |
| LDA | 0.761 | 0.719 | 0.603 | 0.843 | 0.734 |
| QDA | 0.717 | 0.781 | 0.647 | 0.831 | 0.758 |
| SVM | 0.804 | 0.707 | 0.607 | 0.866 | 0.742 |
| Majority Vote | 0.826 | 0.768 | 0.667 | 0.887 | 0.789 |

## 4. Discussion

The results of this study suggest that those with a clinical diagnosis of VVS exhibit significantly different responses to the AS test compared to age-matched CON subjects. In addition, machine-learning approaches were applied successfully to identify a positive clinical diagnosis of VVS. The hemodynamic differences identified allowed the AS to be used to classify those with VVS with sensitivity of 82.6%, specificity of 76.8%, and average accuracy of 78.9%.

The classifier in this study used SBP, HR, SV, CO, TPR, HRV, and BPV features in the identification of subjects with VVS. SBP and HR had been previously used by Mereu et al. (2013) to predict HUTT outcome with 86.2% sensitivity and 89.1% specificity. Differences in our performance metrics are expected given the differences between a HUTT response and clinical diagnosis of VVS.

Ebden (2006) investigated a multivariate LDA classifier on a dataset of 30 patients age 65 and older, using parameters of HRV, BP, HR trend, and instantaneous centre frequency variability (ICFV) taken from the first few minutes after tilting in a HUTT. With this combination of parameters, a PPV of 93%, a NPV of 88%, and an accuracy of 90% were obtained for predicting a positive or negative HUTT result (Ebden 2006). Since the algorithm was tested on older patients, it was expected that similar results might be obtained; however, the data set used by Ebden (2006) was longer in duration, with Ebden analyzing HRV from five minutes of supine rest and upright tilting. Furthermore, this work focussed on predicting HUTT outcomes from early tilting, not to identify a clinical diagnosis.

Virag et al. (2007; 2018) developed a prediction algorithm for predicting HUTT outcome based on calculation of cumulative risk based on the RR and SBP trend, and the LF power of the RR interval (HRV) and the SBP (BPV). The algorithm was tested on a large sample of 1,155 patients and yielded a sensitivity of 95% and specificity of 93% (Virag et al. 2007). This study (Virag et al. 2007) involved a significantly larger dataset allowing for more robust algorithm development; however the prediction times required 23.6 minutes of HUTT data. The algorithm was subsequently evaluated on 140 subjects in a clinical setting and still yielded a sensitivity of 97.6% and specificity 88.2% proving clinical relevance (Virag et al. 2018). Nonetheless it targeted prediction of HUTT outcome before syncope occurred, rather than predicting a clinical diagnosis of VVS.

Carmody et al.’s (2014) algorithm for predicting a clinical diagnosis of VVS in younger adults with the AS test achieved 84.4% sensitivity and 72.97% specificity. However, the features used for classification are somewhat different due to age-related differences in AS responses between younger and older cohorts. For example, the HR percent increase and CO percent increase were found significant for younger adults but not for older adults due to the blunted HR response. This unique hemodynamic response observed in older adults is not surprising. Previously, deviations in the BP and HR responses of healthy older adults compared to healthy younger adults have been observed during the active stand test in the form of slower BP recovery, an initial peak in BP with standing, and a blunted HR increase compared to younger adults (van Wijnen et al. 2017; Wieling et al. 1992; Smith, Porth, and Erickson 1994; Finucane et al. 2019). These responses are characteristic of an aging autonomic nervous system and are evident here.

Furthermore, the differences noted between VVS and age-matched CON in both older and younger cohorts are of interest from a mechanistic perspective. Subjects with VVS demonstrated a smaller initial drop in SBP, a larger decrease in baseline to steady-state values in SV, CO and TPR, and a smaller drop in TPR compared to age-matched CON subjects. VVS subjects had a higher HR throughout the test, as demonstrated by the features in Table 3. Although different in absolute values to the younger cohort, there remain similarities with previous work; i.e. evidence of autonomic hypersensitivity in VVS compared to age-matched controls combined with large drops in SV caused by presumably greater peripheral pooling of blood volume. Carmody et al. had previously identified differences in hemodynamic responses to orthostatic stress in younger adults with VVS compared to healthy younger adults (Carmody et al. 2014). Those with VVS were characterized by lower drops in SBP, and greater overshoot at 20 seconds post-stand, a greater percent increase in HR and CO from baseline, and greater drops in SV from baseline to steady state (Carmody et al. 2014).

### 4.1 Strengths and Limitations

This study used retrospective data. As a result, it was not possible to control for all factors that may influence group differences during data collection or make prospective predictions. The sample size, although the largest group of older adults used to date, was still relatively small and additional subjects would need to be analysed to confirm results in a prospectively designed study. Subtypes of VVS (Vasodepressor and Cardioinhibitory) were not accounted for in the current study and may require additional features to differentiate between subtypes. Since the controls were not clinically assessed for syncope it is was not possible to definitively rule out the presence of VVS in this group. Finally, the AS test analysed during this study is a short test lasting only 2 minutes, is less resource intensive and less stressful for patients making it practical for screening purposes in large populations and cohorts where a HUTT is not feasible or used prior to HUTT. Given previous results involving longer duration tests and higher classification accuracies, it is likely that there is a trade-off between classification accuracy and test duration.

Despite these limitations, the study was the first to focus on an ageing population in the identification of VVS. A broad range of features were investigated and applied as part of a broader machine learning framework to classify a clinical diagnosis of VVS from the AS test. The results indicate the need for further research into the use of the AS test as a screening tool in the diagnosis of VVS and other clinical outcomes.

## 5. Conclusion

Older adults with a clinical diagnosis of VVS exhibit different hemodynamic and autonomic responses to AS characterised by relative autonomic hypersensitivity compared to age-matched controls (with and without VVS). Optimum VVS classification was obtained using a majority vote classifier combining LDA, QDA, and SVM classifiers to achieve 82.6% sensitivity, 76.8% specificity, and 78.9% accuracy. This work highlights the potential value of the AS test as a convenient screening tool for identifying individuals with a clinical diagnosis of VVS. More broadly this paper presents a machine learning framework to support use of the AS to classify clinical outcomes of interest.

## Data Availability

The control subject data was obtained from The Irish Longitudinal Study on Ageing. This data set is lodged in the Irish Social Science Data Archive (www.ucd.ie/issda/data/tilda) and is accessible for bona fide research purposes.
The subject data was obtained from St. James's Hospital, Dublin, Ireland and is unavailable.

## Acknowledgements

The authors would like to thank the patients and staff of the Falls and Syncope Unit at St. James’s Hospital and the participants of the The Irish Longitudinal Study on Ageing (TILDA).

